# Manuscript Title: Facility-based HIV self-testing as an additional testing option in health facilities: A systematic review and meta-analysis

**DOI:** 10.1101/2024.04.19.24305307

**Authors:** Kathleen McGee, Muhammad S. Jamil, Nandi Siegfried, Busisiwe Msimanga Radebe, Magdalena Barr-DiChiara, Rachel Baggaley, Cheryl Johnson

## Abstract

Facility-based HIV self-testing (FB-HIVST) has been used across settings to improve testing accessibility and achieve global testing and treatment targets by 2030. The effectiveness of FB-HIVST remains uncertain; thus, we conducted a review to assess the risk and benefits of FB-HIVST to inform global guidance. We searched across nine electronic databases covering the period up to February 01, 2022, and included publications that directly compared FB-HIVST to standard HIV testing services (SOC) or no intervention. Meta-analysis was conducted on comparable outcomes using random-effects model for relative risks (RR) and 95% confidence intervals. Other outcomes were summarized descriptively. Risk of bias was assessed using Cochrane’s Risk of Bias tool. Certainty of evidence was assessed using Grading of Recommendations, Assessment, Development and Evaluations (GRADE). After screening 2,203 articles, 11 studies were found eligible, including 4 randomized controlled trials (RCT), 2 cohort studies, 3 economic evaluations, and 2 qualitative studies. Meta-analyses of four RCTs demonstrated that FB-HIVST may increase testing uptake (RR=2.47; 95% CI= 0.96, 6.33) and may lead to greater HIV diagnosis (RR=3.77; 95% CI=0.81, 17.44). Overall GRADE certainty was low. Trials found FB-HIVST as acceptable and feasible to many users, with minimal risk of social harm. A single RCT reported on linkage to care and observed that, among total enrolled, FB-HIVST compared to SOC may increase linkage to care threefold (RR= 3.26; 95% CI: 0.68, 15.62; low-certainty evidence). FB-HIVST was found to be cost-effective in a high-burden outpatient department, but determined to be quite variable. FB-HIVST is safe and may be an effective method to increase testing coverage and the diagnoses, particularly in high-burden HIV settings or sites with limited staff and resources. Findings from this review informed WHO’S guideline development process and its recommendation that FB-HIVST be offered as an additional testing option at facilities.

PROSPERO Number: CRD42022302619

## Introduction

Since 2016, the World Health Organization (WHO) has recommended HIV self-testing (HIVST) as a safe and effective HIV testing option [1], whereby an individual collects their own specimen, using a rapid finger-prick or oral fluid-based test, performs the test, and interprets their result, with or without provider assistance. Since then, the range of WHO-prequalified HIVST products has expanded [2], and over 100 countries have integrated HIVST into their national policies and programmes [3]. Many different service delivery models for HIVST have been rolled out worldwide to better reach people with HIV who do not know their status, as well as to make testing easy for people with ongoing HIV risks. Numerous self-testing approaches have been shown to be effective testing strategies, such as distribution in communities, workplaces, faith-based settings, online via mail, retail outlets, pharmacies, vending machines, and secondary distribution to partners or peers [4–6].

In high-burden HIV settings, distribution of HIVST kits in health facilities has been used to increase testing coverage [4]. During the COVID-19 pandemic, many health facilities offered HIVST to maintain testing services while mitigating added strain on limited human resources [5,6]. Prior to the pandemic, some programmes began to use risk-screening tools among adults and adolescents to address limited resources and focus HIV testing to specific risk factors [7]. Risk screening tools have been especially explored for provider-initiated testing and counselling (PITC) in east and southern Africa. Despite the potential for optimizing resources by targeting testing, recent evidence suggests they may have contributed to missing cases and undermine global HIV goals [8]. To date, WHO does not recommend the use of “screen-out” tools that are designed to exclude low-risk people from routine HTS. However, a recent systematic review suggests that “screen-in” risk-based tools may be useful in settings where the routine offer of HTS is not currently available or feasible and could be further optimised with facility-based HIVST (FB-HIVST) [7].

As programmes seek to optimise increasingly limited resources for HIV testing services, policy makers and implementers are grappling with whether FB-HIVST should be considered [9]. To support the World Health Organization (WHO) in updating existing HIVST guidance and understand the potential benefits and risks of FB-HIVST uses, we conducted a systematic review of the literature. This review informed the 2023 consolidated guidelines on HIV testing services.

## Materials and Methods

The review protocol was registered online in the International prospective register of systematic reviews (PROSPERO 2022 CRD42022302619).

### Search strategy and inclusion criteria

We searched nine electronic databases including Ovid Medline, Embase, CINAHL Plus, EconLit, Global Health, PsycInfo, Cochrane Library, Web of Science, and Scopus through January 2022. We also verified secondary references on studies included in the review as well as on previously published review articles relevant to HIVST. Experts in the field were contacted to identify additional articles and validate inclusion criteria. As previously stated, this paper presents the results as they were presented to WHO’s guideline development group in November 2022 leading to WHO guideline recommendations in July 2023.

The search strategy was adapted to each database using key terms “HIV”, “self-test” (Appendix 1, Supplementary material). HIVST in any type of health facility setting were eligible, with the exception of secondary distribution. No restrictions were placed based on study location or language of the article. Two reviewers screened studies and differences in judgement were resolved by other review team members. To be included in the review, a study had to directly compare FB-HIVST to standard HIV testing services (HTS), or no intervention, and report on one or more of the following outcomes: (1) HIV testing uptake (defined as proportion of participants tested for HIV in study period); (2) HIV positivity (proportion of people tested and confirmed positive); (3) linkage to care (proportion of people tested positive linked to confirmatory testing, clinical assessment or treatment); (4) social harm/adverse events (occurrence and reporting of any harm or undesirable experience occasioned by the intervention); (5) acceptability of FB-HIVST (proportion of participants who reported intervention as acceptable); (6) diagnostic accuracy (measured by specificity, sensitivity, positive predictive values, and negative predictive values); (7) Feasibility (proportion of participants who reported intervention as easy to use); (8) values and preferences (reported facilitators and barriers associated with the intervention), (9) costs and cost-effectiveness.

### Data analysis

Data was extracted by one reviewer using an Excel-based data extraction form. Risk of bias was assessed according to Cochrane Guidance [10]. Pair-wise meta-analysis was conducted on comparable outcomes, when possible, using REVMAN 5.4.1, using random-effects model for relative risk (RR), with 95% confidence intervals (CI). Following guidance from GRADE Handbook [11], overall certainty of evidence was rated by appraising risk of bias, level of imprecision, indirectness, inconsistency and other considerations. In accordance with WHO guideline development protocol, we report the results using GRADE recommended language which integrates the clinical importance of the estimate of effect with the overall certainty of the estimate [12]. Evidence relating to values and preferences such as perceived benefits and barriers, reasons for uptake or choice of testing, and participant recommendations were summarized qualitatively. Full RoB and GRADE Quality Assessment is available in (Supplementary material).

## Results

The searches yielded 2,203 citations, of which, 11 studies were deemed eligible, including 4 randomized controlled trials [4,13–15], 2 prospective single-arm cohort studies [16,17], 2 qualitative studies employing individually-based in-depth interviews (IDI) [18,19], 2 costing studies [20,21] and 1 cost-effective analysis [22] (Figure 1).

**Figure 1.**
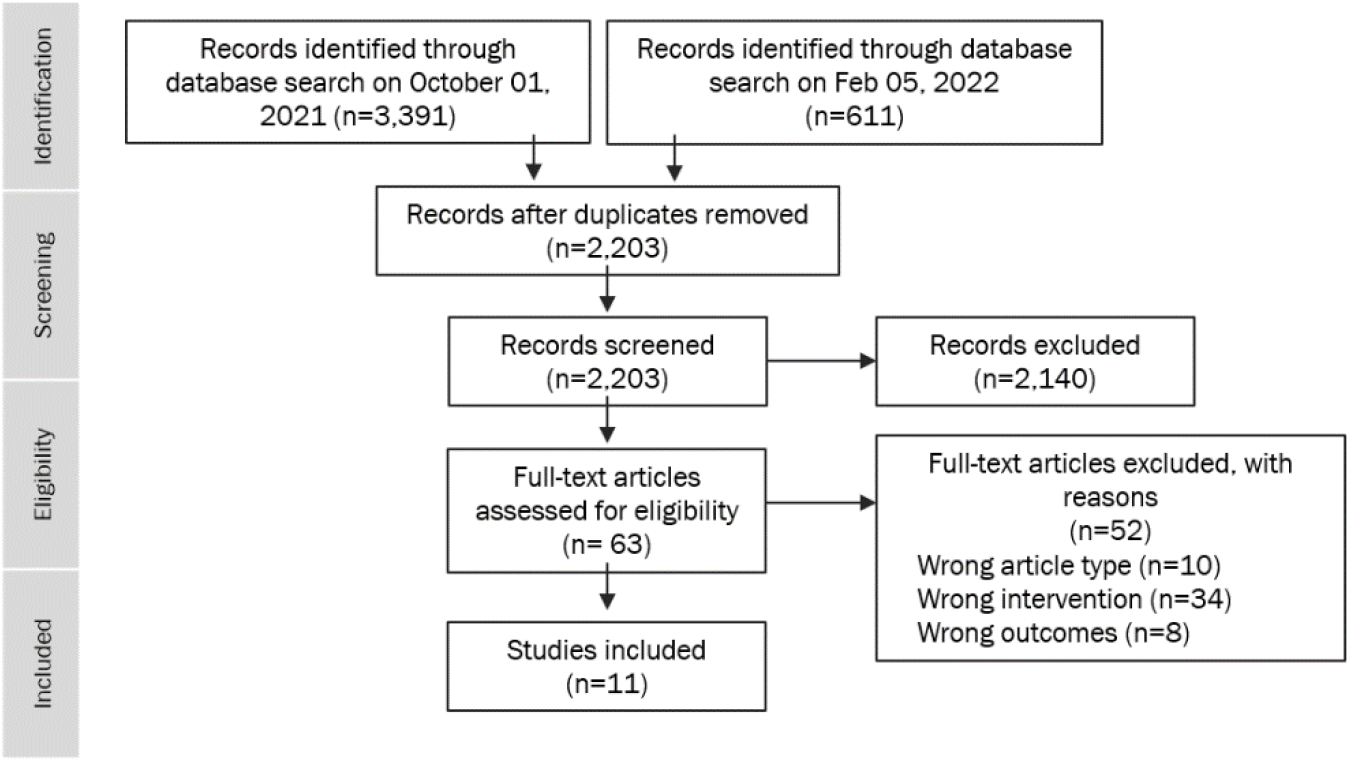
Prisma Diagram.

Among the 4 RCTs, Dovel 2020 [4], a cluster RCT, served as a parent trial for subsequent studies in Malawi, including a costing study [20], a modelled cost-effectiveness study [22], and a qualitative study [18], targeting adolescent & adult outpatients in high burden health facilities. The remaining RCTs [13– 15] were individual RCTs, targeting truck drivers and female sex workers at roadside wellness clinics in Kenya. Additional studies were set in a U.S. emergency department (ED) [16], youth-friendly hospitals in Mozambique [17], health facilities in Malawi [19], and ante-natal clinics (ANC) and out-patient departments (OPD) in Zambia and Zimbabwe [21], targeting adult and adolescent outpatients. All studies used oral fluid-based HIV rapid diagnostic tests for self-testing, providing participants with free test kits and manufacturer-provided instructions. See Table 1. for further information on studies’ implementation details.

**Table 1.**
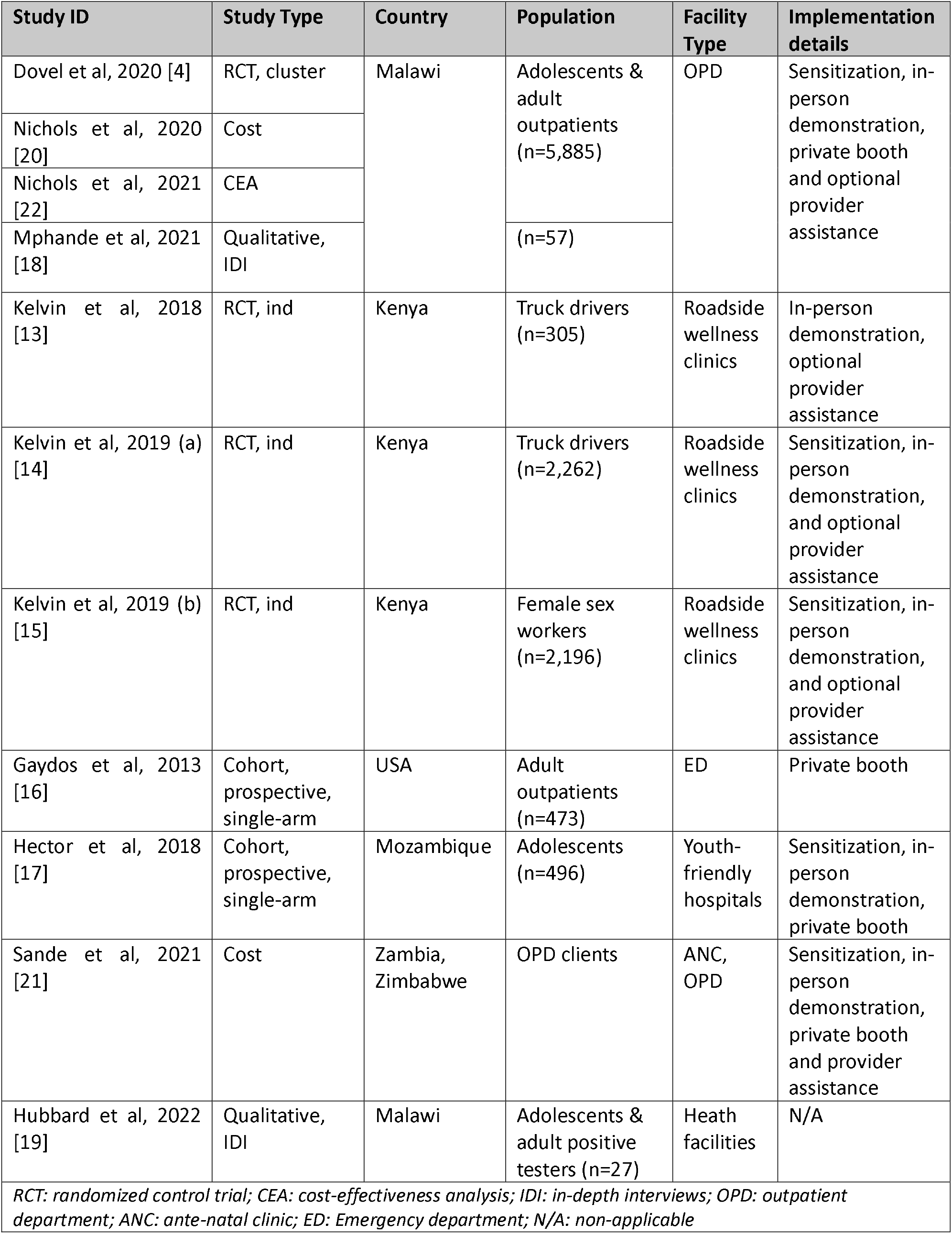
Summary of included study characteristics (n=11)

### HIV Testing Uptake

Four RCTs [4,13–15] reported on HIV testing uptake. A meta-analysis showed that FB-HIVST may improve HIV testing uptake compared to standard HTS (RR= 2.47; 95% CI: 0.96, 6.33; Chi^2^ = 187.21; df = 3; p<0.00001; I^2^ = 98%, low certainty evidence). In absolute numbers, there were over twice as many testers in the FB-HIVST arms compared to testers in the standard of care (SOC) arm.

**Figure 1.**
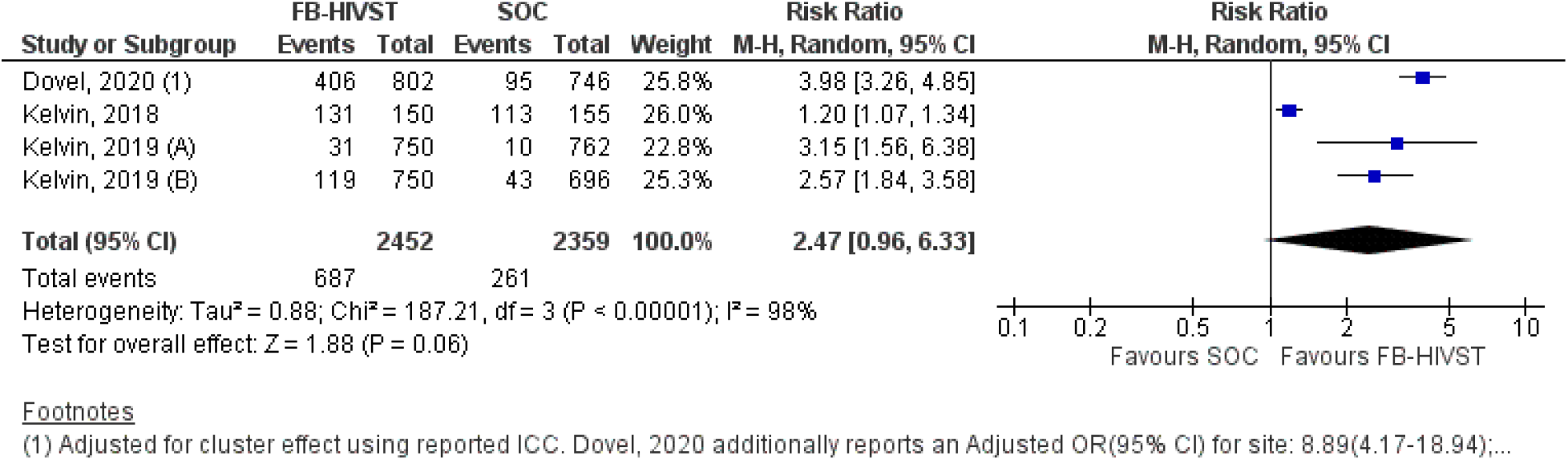
HIV testing uptake among participants offered FB-HIVST vs. HTS. FB-HIVST: Facility-based HIV self-testing; CI: Confidence Interval; SOC: standard of care; M-H: Mantel-Haenszel; OR: odds ratio

The overall certainty was rated as low due to a high risk of bias combined with the non-statistical significance of the pooled effect. High levels of statistical heterogeneity were driven by Kelvin, 2018, where the study population did not receive any sensitization about HIV testing or HIVST (Appendix 2, Supplementary materials). In the three other RCTs, sensitization included a 10-minute health talk to all outpatients present in the waiting room, and text messages on the importance of HIV testing.

The test for subgroup differences between population types didn’t present a statistically significant subgroup effect. Rather, subgroup analysis showed FB-HIVST may increase HIV testing uptake among both the general population and key population groups, compared to standard testing alone. In a single RCT conducted among a general population [4] FB-HIVST resulted in nearly a four-fold increase in HIV testing uptake compared to standard testing (RR= 3.98; 95% CI: 3.26-4.85, low-certainty evidence). Subgroup analysis of three RCTs [13–15] conducted among key and priority populations, determined that FB-HIVST could potentially double testing uptake compared to standard testing, however the effects are uncertain due to very low-certainty evidence (RR= 2.07; 95% CI: 0.86, 4.97; Chi2 = 43.63; df = 2; p<0.00001; I2 = 95%).

### HIV Positivity

All four RCTs reported on HIV positivity [4,13–15]. A meta-analysis of these studies determined FB-HIVST may likely diagnose more people with HIV compared to standard HTS (RR= 3.77; 95% CI: 0.81,17.44; Chi^2^ = 4.67; df =3 ; p<0.20; I^2^ = 36%; low-certainty evidence). Certainty was again downgraded to low due to high risk of bias and a large confidence interval crossing the line of no effect, thus driving imprecision. In absolute numbers, FB-HIVST diagnosed more than five times as many people with HIV than standard HTS. Observed heterogeneity was driven by Kelvin 2018 [13], which, unlike other studies, did not provide any sensitization on HIV testing.

**Figure 2.**
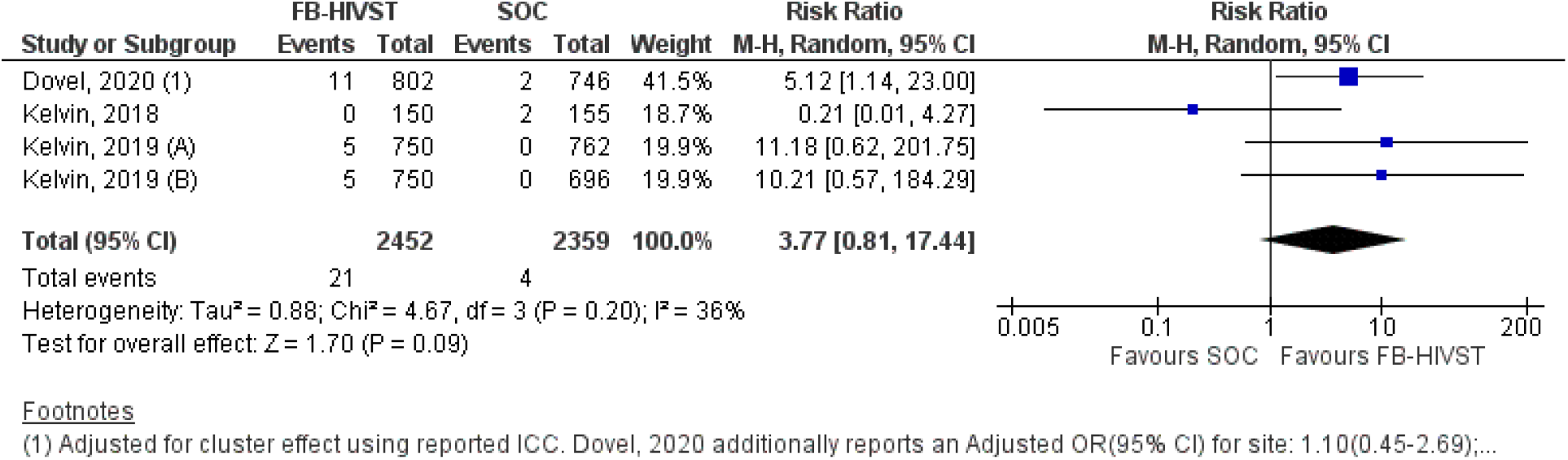
HIV Positivity. FB-HIVST: Facility-based HIV self-testing; CI: Confidence Interval; SOC: standard of care; M-H: Mantel-Haenszel; OR: odds ratio

Subgroup analysis suggests that, when compared to standard HTS, there is very low-certainty evidence that FB-HIVST plus sensitization could potentially increase the number people with HIV diagnosed (Appendix 2, Supplementary materials). HIV positivity could also be higher when using FB-HIVST, instead of standard HTS alone, in the general outpatient population in a high HIV burden setting (RR= 5.12; 95% CI: 1.14, 23.00). However, FB-HIVST could make little to no difference on HIV positivity in clinics reaching key and priority populations when compared to standard testing (RR= 2.97; 95% CI: 0.23, 37.65 ; Chi^2^ = 4.48; df = 2; p<0.11; I^2^ = 55%).

Other subgroup analysis revealed no other clear factors driving heterogeneity (Appendix 2, Supplementary materials).

### Linkage to HIV Care

Only one RCT [4] reported on linkage to care and reported that FB-HIVST, compared to standard HTS, may increase the number of people linked to HIV care after three-months three-fold (RR= 3.26; 95% CI: 0.68, 15.62). This effect was driven by the high number of people tested and diagnosed with HIV in the study which took place in an outpatient department. When the analysis was limited to those newly diagnosed with HIV, however, there was no difference in linkage rates between FB-HIVST and standard HTS (RR= 0.81; 95% CI: 0.52-1.26) (Appendix 2, Supplementary materials). Overall, there was low-certainty evidence that FB-HIVST could enable more people with HIV to link to care overall or at least achieve linkage rates comparable to standard testing services.

### Social harm

Social harm was only reported in one RCT [4] which found harms to be rare, particularly for FB-HIVST compared to SOC. In this study participants were asked about harms when testing, including if they experienced coercion or pressure to test and disclose results. More participants reported that they experienced pressure to test (n=10) or to share their test results (n=1) in provider testing, while no participants receiving FB-HIVST reported experiencing such pressure. Thus, there is low-certainty evidence that FB-HIVST does not increase social harm and may even result in fewer harms compared to standard facility-based testing.

### Acceptability

Acceptability of FB-HIVST was reported by all four RCTs [4,13–15], and all studies found that, when compared to standard testing, most individuals would probably prefer FB-HIVST (moderate-certainty evidence).

In one study among individuals in general outpatient departments [4], those receiving FB-HIVST were more likely to test again (RR= 1.21; 95% CI: 1.10-1.33) and more likely to recommend testing to friends (RR= 1.12; 95% CI: 1.04-1.21) compared to standard facility-based HTS. In RCTs among key (female sex workers) and vulnerable populations (long-distance truck drivers) [13–15], FB-HIVST was also slightly more preferred when compared with both home-based HIVST and standard testing (FB-HIVST: 16.8%, 151/900, home-based HIVST: 4.0%, 36/900, standard testing: 10.3%, 93/900).

### Feasibility and diagnostic accuracy

No RCTs reported on feasibility and accuracy related outcomes. Thus, findings were derived from two observational cohort studies [16,17]. With 75.33% (n=577/766) of pooled participants reporting HIVST easy to use, both studies found that adolescent and adult users (16 to 64 years old) generally found FB-HIVST feasible [16,17]. However ease of use appeared highest among adults in emergency departments in the USA (99%, n=463/467) [16].

Only one study reported on the accuracy and performance of FB-HIVST compared to confirmatory finger-prick HIV test [17] The study reported acceptable sensitivity (100%, 95%CI: 99-100%) and specificity (100%, 95%CI: 48-100%). No false negative or false positive results were identified, but confidence intervals around specificity were particularly wide due to small numbers. Additionally, there were six reports of inconclusive or invalid results identified: two nonreactive self-test results which were reported as inconclusive following provider-delivered testing and four invalid self-tests which were either confirmed negative (3) or were inconclusive (1) following provider-delivered testing. The study did not specify whether invalid testers were already on anti-retroviral treatment.

### Values and preferences

Five studies, including 2 RCTs [4,13], 1 cohort [17], and 2 qualitative studies [18,19], reported on some aspect of user values and preferences for FB-HIVST among adult outpatients, truckers, adolescents between ages 16-20, and men who tested HIV positive. In all studies, most reported that FB-HIVST was acceptable and feasible and addressed some concerns about testing in clinics.

Five studies reported on user values and preferences for FB-HIVST [4,13,17–19]. In all studies, most reported that FB-HIVST was acceptable and feasible and addressed some concerns about testing in clinics. Continuing to receive demonstrations on how to self-test and tailored post-test services as part of FB-HIVST remained important to many [4,19].

Common factors for preferring facility-based HIVST included a desire for oral fluid-based testing, interest in trying something new, greater convenience, autonomy and easy access to in-person support if needed. Among adolescent testers in Mozambique [17], 85% (n=253) said they preferred FB-HIVST over the standard HTS with most wanting FB-HIVST because it didn’t require a finger prick (56.4%) and enabled access to counselling and privacy from family (76.3%). Truck drivers who opted into FB-HIVST had some similar reasons, with many preferring oral testing (25%), and feeling curious to try something new (89%) [13]. In the general outpatient population, however, most valued FB-HIVST for its speed, convenience, autonomy and easy access to post-test services [4]. Some also felt more motivated to test because they saw others self-testing and that felt that if they had not self-tested at the facility they may not have linked to care [19].

Not all individuals preferred FB-HIVST. For those desiring standard testing services, many did not trust oral fluid-based tests or felt they would not be able to self-test correctly [17]. Others, however, preferred to self-test at home instead of at a facility [17], so they could test with a partner, family member or friend [13], and could avoid unwanted disclosure or judgement at facilities [18].

### Costs and Cost-Effectiveness

Three studies reported on resource use [20–22] and were all in sub-Saharan Africa (see summary in Appendix 3, Supplementary materials). In one multi-country study [21], the average cost per HIVST kit distributed in facilities was less than or equal to home-based HIVST. And in a costing study which used data from a RCT [20], FB-HIVST had a higher average incremental cost per person tested, per person newly diagnosed, and per patient initiated on ART, than standard HTS. However, when results were adapted and modelled to public health clinics, the average unit costs for FB-HIVST was more comparable to standard testing. Despite somewhat higher costs than standard facility-based HTS, when using this same data a modelling study found that when assuming a threshold analysis of $200 USD (2017) per new diagnosis, FB-HIVST would be cost-effective and potentially cost-saving if lower cost HIVST kits ($1.80-1.00) were used or if FB-HIVST was limited to individuals who had not tested in the past year [22].

Consumables and staffing costs were the main cost drivers of FB-HIVST. However, time and motion data from one study [4] did identify that FB-HIVST be used to reduce provider time by 36-53% and thereby lead to cost-savings in facilities.

## Discussion

HIVST has become a critical tool for maintaining essential services and expanding access to testing, prevention, and care. This systematic review and meta-analysis indicate low certainty evidence that FB-HIVST, compared to standard testing, increases testing uptake and diagnoses, proving acceptable and potentially affordable, especially in low-income countries. These findings align with previous reviews on the effectiveness of HIVST across settings, populations, and implementation strategies.

FB-HIVST has demonstrated potential in promoting testing uptake and diagnosis. Amidst the challenges posed by the COVID-19 pandemic, FB-HIVST emerged as an effective tool, showcasing its relevance for future pandemic preparedness and addressing human resource-constraints [5,6]. The results from this review further signal an opportunity for FB-HIVST to replace risk-based screening tools, for which, recent data suggests may inadvertently reduce testing coverage, diagnosis, and ART initiations [8]. FB-HIVST would offer a streamlined and more accessible alternative, especially in settings with complex risk-screening protocols.

Additionally, the systematic review found that FB-HIVST affords users the customary benefits associated with self-testing, such as convenience, autonomy, and ease of use, while also providing the added advantage of integrating post-test facility-based services. Notably, a more recent qualitative study has also found that FB-HIVST may help overcome traditional facility-based barriers to HIV testing [23]. Analysis on acceptability reveals that individuals expressing a preference for FB-HIVST exhibit an inclination towards oral testing. It is noteworthy, that not all HIV self-tests are oral, and existing literature highlights the acceptance of blood-based self-tests [24–26]. Optimizing FB-HIVST implementation requires strategies for private and convenient service delivery, such as provision of private self-testing spaces [23]. The significance of demonstrations and sensitization in enhancing user experience should also leveraged, acknowledging that various HIVST methods remain essential to cater to diverse preferences and needs.

Linkage to care evidence was drawn from a single trial. On one hand, linkage to care among total participants enrolled in the study, was over three times as likely in FB-HIVST than standard HTS, driven by the greater number of people diagnosed with HIV through FB-HIVST. On the other hand, linkage to care rates among people newly diagnosed with HIV may be lesser in FB-HIVST than standard HTS. In other words, the interpretation of linkage to care may vary depending on the analysis conducted. In short, despite FB-HIVST’s potential to diagnose and link more people to care overall, it still introduces a risk of loss-to-follow-up as linkage to care is contingent on self-reporting. FB-HIVST may thus necessitate follow-up strategies such as providing HIVST kits with linkage to care information, and in-person or remote post-test counselling services [30]. Notably, no study reported on linkage to prevention services. Given the new recommendation to integrate HIVST to Pre-Exposure Prophylaxis (PrEP) services [31], it is important for future studies to further examine strategies for linkage to prevention services.

Only one cohort study compared the diagnostic accuracy of FB-HIVST to the results of confirmatory finger-prick tests. Overall accuracy was high, though there were some occurrences of diagnostic discrepancies (2%, n=6/299). Other studies evaluating user’s interpretations of HIVST results have similarly found users less likely to correctly interpret invalid and weak positive results, both for oral fluid and blood-based/finger-prick HIVST [32,33]. In the case of FB-HIVST, the potential to support clients in their interpretation of HIVST may be more readily available in the facility. Nevertheless, sensitization and demonstrations, as well as written and visual aids should be further explored to support all HIVST clients. Furthermore, the increasing use of videos and alternative instructional methods, informed by lessons learned during the COVID-19 pandemic, hold promise for enhancing HIVST accuracy.

FB-HIVST extends beyond HIV diagnostics, potentially contributing to broader self-care and self-testing models. As evidenced by emerging options such as the dual HIV/Syphilis self-test kits, FB-HIVST sets a precedent for integrating multiple disease screenings in a single, unified step. This innovation suggests potential applications across other disease programs, with opportunities for adaptation to areas such as COVID-19, Hepatitis C infection, and other sexually transmitted infections. The scalability and adaptability of FB-HIVST may present an exciting prospect for streamlining diagnostic processes, enhancing accessibility, and ultimately improving greater public health outcomes.

In regard to costs, the availability of a $1 USD HIVST listed by WHO prequalification [34] marks progress in affordability and accessibility. While early evidence signals FB-HIVST may help save health worker time, it is important to further investigate the full costs of this approach in real-world settings, including in cases where pre-testing demonstrations and optional support to self-test users are provided. Understanding how time and resources can be adjusted to maximize potential savings is crucial to support effective use of this testing approach. Finally, costs of FB-HIVST across low and high-burden settings may be further investigated to ascertain whether the intervention remains cost-effective across different settings.

This review was limited by the number of trials which directly compared FB-HIVST to standard HTS so that meta-analysis could only be performed for two reported outcomes. Certainty of evidence was commonly downgraded to low due to high risk of bias and imprecision, driven by confidence intervals encompassing the null effect. The studies were primarily conducted in east and southern Africa and may thus not be generalizable to other settings. Further research is needed to evaluate whether the costs and benefits of FB-HIVST hold true in low-burden settings, where progress toward testing and treatment targets may lag. This exploration is crucial, particularly in contexts where FB-HIVST could offer new testing opportunities to underserved populations, such as men and adolescents. Likewise, while information on resource use was identified it was very limited.

Based on the findings of this review, coupled with additional evidence reviewed at an expert meeting, WHO recommended in July 2023, that FB-HIVST be offered as an additional option for testing in facilities [31]. Ultimately, HIVST can continue to play an important role in increasing access to HIV testing services, driving case-finding efforts to reach those who have not yet benefitted from HIV testing, and promoting efficiency in resource-constrained environments.

## Conclusion

This review found FB-HIVST may be a safe, acceptable, and effective method to increase HIV testing coverage and diagnosis. These findings are particularly relevant for clinical settings with high HIV burden and limited staff and resources and where there continues to be gaps in HIV testing service delivery. Based on these findings, WHO now recommends FB-HIVST be offered as an additional testing option in facilities. HIVST does not replace provider-administered testing for those with reactive results. Individuals with a reactive self-test result should receive further testing from a trained provider using the full national testing algorithm. Further research on ways to optimise service delivery, linkage to prevention and care and allocation of limited resources remain a priority.

## Competing interests

We declare no competing or conflict of interest.

## Authors’ contribution

CJ and MSJ conceived and provided overall guidance to review. KVM conducted screening, data extraction, analysis, with support from CJ, MSJ, and NS. KVM and CJ drafted the manuscript, and all authors MSJ, NS, BMR, MBDC, RB, and CJ contributed, read, and approved the final version.

## Acknowledgements

We would like to thank Margaret Crampton for her assistance with the screening stage of this review as well as the World Health Organization guideline development group members for their contributions.

## Funding

This systematic review was supported by Unitaid-WHO HIV and Co-Infections/Co-Morbidities Enabler Grant [HIV&COIMS], 541 no grant number; https://unitaid.org)

## Data Availability Statement

The data extracted for this systematic review are available within the article and its supplementary materials.

## Notes

### Competing Interest Statement

The authors have declared no competing interest.

### Clinical Protocols

https://www.crd.york.ac.uk/prospero/display_record.php?RecordID=302619

