## Supplementary Information for "Manuscript Title: Facility-based HIV self-testing as an additional testing option in health facilities: A systematic review and meta-analysis"

**Supplementary Materials**

**Appendix 1. Search Strategy**

| Ovid Medline | |
| --- | --- |
| Action | **Term** |
| 1 | exp HIV Infections/ |
| 2 | exp HIV/ |
| 3 | hiv.ti,ab |
| 4 | "hiv1".ti,ab. |
| 5 | "hiv2".ti,ab |
| 6 | “hiv type 1".ti,ab. |
| 7 | "hiv type 2".ti,ab. |
| 8 | human immunodeficiency virus.ti,ab |
| 9 | human immunedeficiency virus.ti,ab. |
| 10 | human immuno-deficiency virus.ti,ab |
| 11 | human immune-deficiency virus.ti,ab |
| 12 | (human immun* adj3 deficiency virus).ti,ab |
| 13 | acquired immunodeficiency syndrome.ti,ab |
| 14 | acquired immunedeficiency syndrome.ti,ab |
| 15 | acquired immuno-deficiency syndrome.ti,ab |
| 16 | acquired immune-deficiency syndrome.ti,ab |
| 17 | (acquired immun* adj3 deficiency syndrome).ti,ab |
| 18 | Sexually Transmitted Diseases, Viral/ |
| 19 | or/1-18 |
| 20 | (sample adj1 collect*).ti,ab |
| 21 | Dried Blood Spot Testing/ and (home* or remote or personal or self*).ti,ab |
| 22 | (dried blood spot adj3 (home* or remote or personal or self*)).ti,ab |
| 23 | (alternative adj3 test*).ti,ab |
| 24 | (option* adj1 test*).ti,ab |
| 25 | ((Home* or self* or mail*) adj3 (collect* or sampl* or specimen* or test* or kit)).ti,ab |
| 26 | or/20-25 |
| 27 | 19 and 26 |

**Appendix 2. Sub-group analysis**

#### *.1 Subgroup analysis: HIV testing uptake*

Figure A4.1.1. HIV testing uptake in populations sensitized to the importance of HIV testing, and among populations who did not receive sensitization prior to intervention.

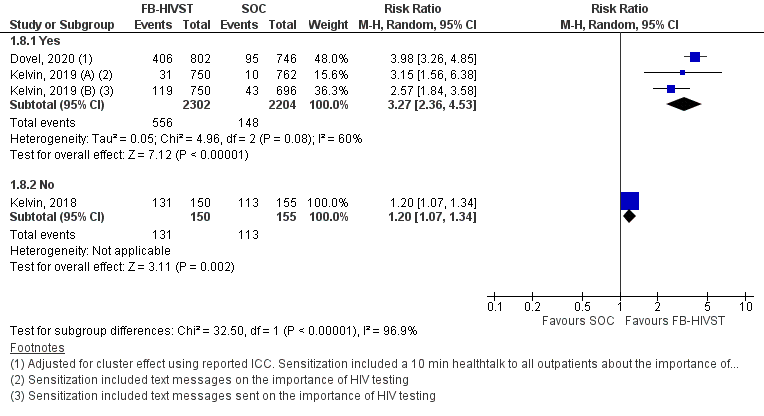

Figure A2.1.2. HIV testing uptake in general populations and in key and priority populations.

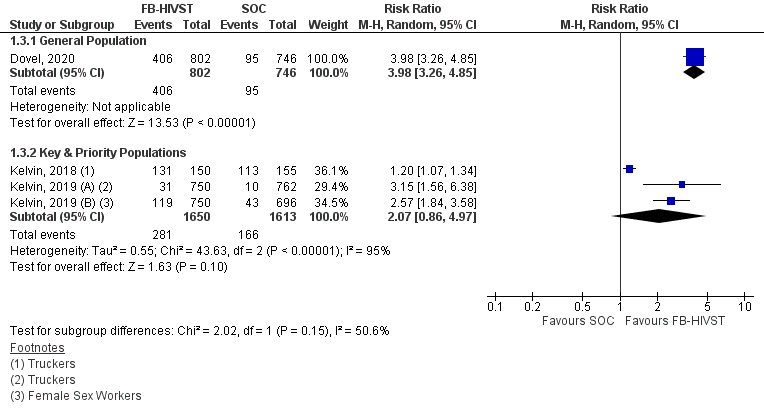

Figure A2.1.3. HIV testing uptake in facility-based HIVST vs. standard of care, and facility-based HIVST vs. enhanced standard of care.

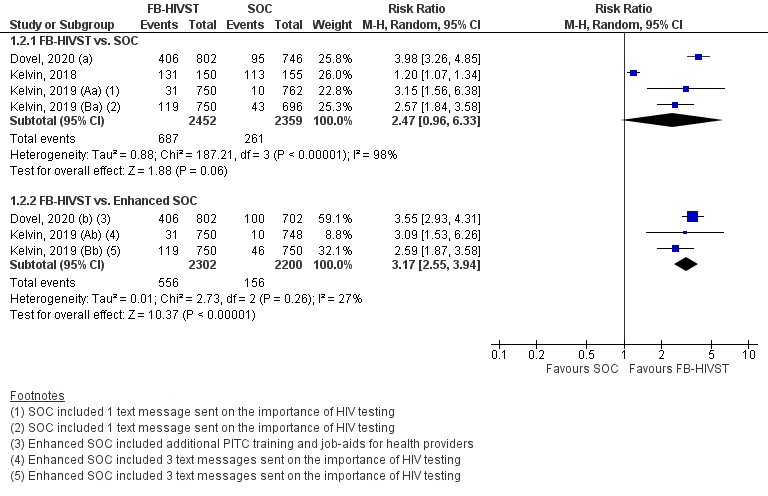

Figure A2.1.4. HIV testing uptake among participants provided with private booths for facility-based HIVST, and participants who were not provided with private booths for facility-based HIVST.

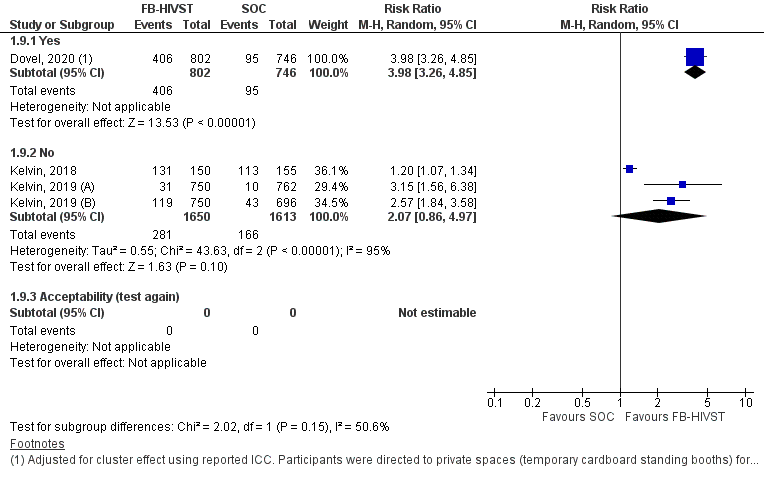

#### *2.2 Subgroup analysis: HIV positivity*

Figure A2.2.1. HIV positivity in populations sensitized to the importance of HIV testing, and among populations who did not receive sensitization prior to intervention.

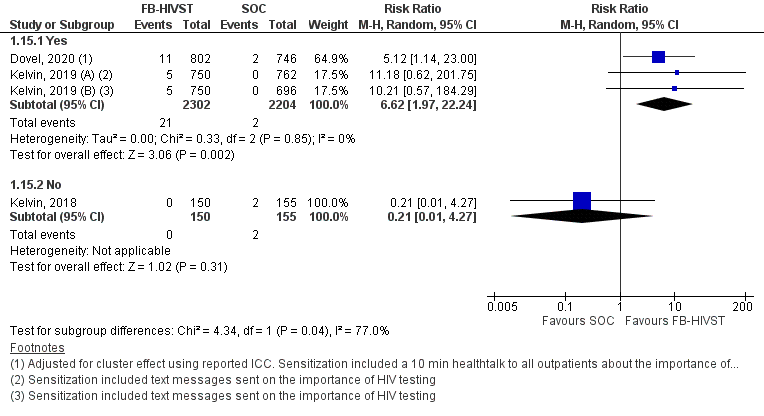

Figure A2.2.2. HIV positivity in general populations and in key and priority populations.

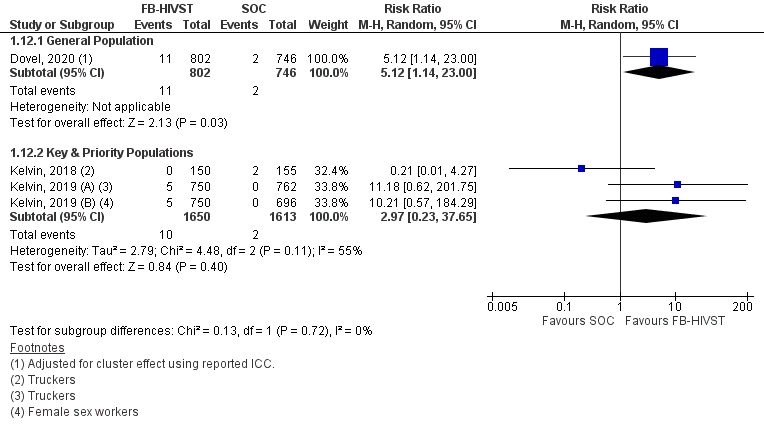

Figure A2.2.3 HIV positivity in facility-based HIVST vs. standard of care, and facility-based HIVST vs. enhanced standard of care.

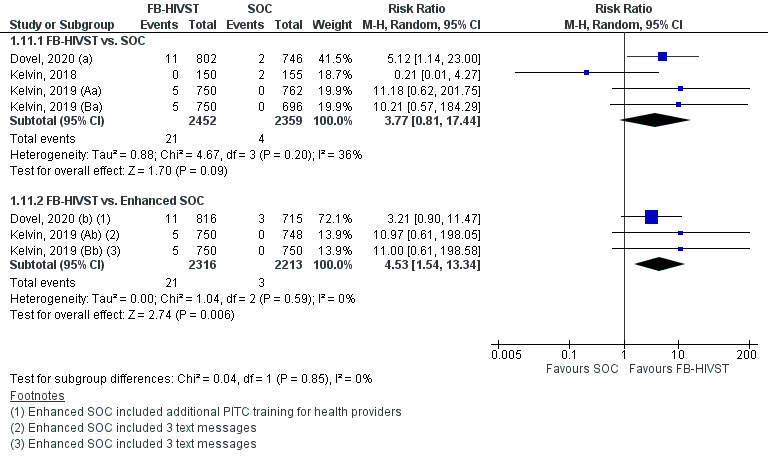

**Appendix 3. Summary of reported unit costs, in 2021 USD**

| **Study** | **Country** | **Study Type** | **Cost Type** | **Yr** | **Cost per HIVST kit distributed** | | **Cost per person tested** | | **Cost per new HIV diagnosis** | | **Cost per patient initiated on ART** | |
| --- | --- | --- | --- | --- | --- | --- | --- | --- | --- | --- | --- | --- |
|  | | | | | **FB-HIVST** | Home-HIVST | **FB-HIVST** | PITC | **FB-HIVST** | PITC | **FB-HIVST** | PITC |
| Nichols, 2020 | Malawi | Obs | Fin, Inc | 2021 | - | - | **$6.63** | $3.24 | **$251.25** | $134.26 | **$384.18** | $160.85 |
|  |  | Mod ^†^ | Fin, Inc | 2021 | - | - | **$3.26** | $3.24 | **$123.63** | $134.26 | **$182.12** | $160.85 |
| Nichols, 2021 | Malawi | Mod | Fin, Inc | 2021 | - | - | **$2.29** ^‡^ | $1.77 | - |  | - |  |
| Sande, 2021 | Zambia | Obs | Econ, Inc | 2021 | **$3.88** | **$5.87** | - |  | - |  | - |  |
|  | Zimbabwe | Obs | Econ, Inc | 2021 | **$22.84** | **$22.37** | - |  | - |  | - |  |
| ^†^ In modelled routine scenario, study staff salaries replaced with country MOH salaries + annual community sensitization costs  ^‡^ Unit cost per person tested calculated from weighted average between cost of person tested positive and negative using study’s assumed testing yield of 2.5%  USD costs converted to 2021 USD, as demonstrated in Kumaranayake, 2000. Note that unit costs converted to 2021 USD in Zimbabwe appear very high due to country hyperinflation.  Obs: Observed study; Mod: Modelled; Fin: Financial; Econ: Economic; Inc: Incremental | | | | | | | | | | | | |

**Appendix 4. Risk of Bias Assessment**

**4.1 Cochrane Risk of Bias Assessment for Randomized Controlled trials**

Figure 4.1.1. Risk of bias graph: review of authors' judgements about each risk of bias item presented as percentages across all included RCTs

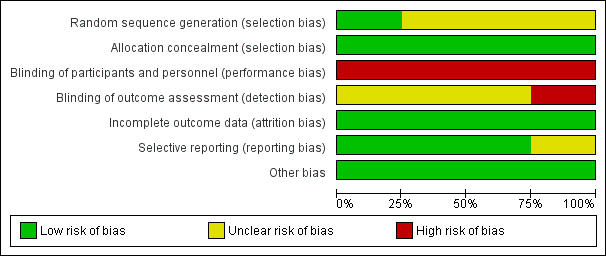

Figure 4.1.2 Risk of bias summary: review of authors’ judgements about each risk of bias item for each included RCT

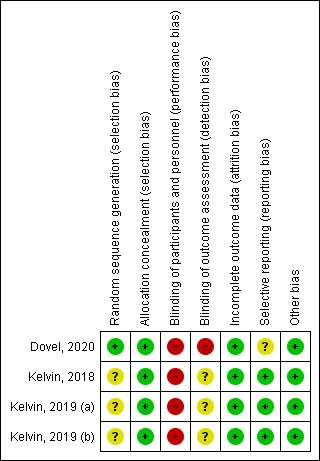

Table 4.1.1 Cochrane risk of bias table by RCT

| **Author/Year** | **Dovel et al, 2020** |
| --- | --- |
| Random sequence generation (selection bias) | **Low risk**  Random assignment was done by statistician using a randomized computer-generated sequence |
| Allocation concealment  (selection bias) | **Low risk**  Trial was clustered at facility level. Outputs were shared with ministry of health, district-level governance and medical providers at participating sites. |
| Blinding of participants and personnel (performance bias) | **High risk**  Neither participants nor personnel could be blinded to intervention. |
| Blinding of outcome assessment (detection bias) | **High risk** (for uptake, positivity, acceptability, and social harm)  Outcomes were self-reported. Authors also note that there may be risk of unmeasured biases resulting from study sampling method.  **Low risk** (for linkage to care)  Outcome was measured from review of clinical records at participating and surrounding facilities. |
| Incomplete outcome data  (attrition bias) | **Low risk**  Less than 2% attrition |
| Selective reporting  (reporting bias) | **Unclear risk**  Not all outcomes specified in trial registry were reported in manuscript, e.g.: HTS outcomes in partners. |
| Other bias | **Low risk** |

| **Author/Year** | **Kelvin et al, 2018** |
| --- | --- |
| Random sequence generation (selection bias) | **Unclear risk**  Randomized but method not specified. Distribution of baseline patient characteristics suggest randomization was achieved. |
| Allocation concealment  (selection bias) | **Low risk**  Participants were not informed they would be randomized to different arms so to avoid selection bias. |
| Blinding of participants and personnel (performance bias) | **High risk**  Participants nor personnel were blinded |
| Blinding of outcome assessment (detection bias) | **Unclear risk**  Unclear if outcome assessment came from self-report or facility records and if this was done equally across arms. |
| Incomplete outcome data  (attrition bias) | **Low risk**  No evidence of loss-to-follow-up |
| Selective reporting  (reporting bias) | **Low risk**  No evidence of selective reporting |
| Other bias | **Low risk** |

| Author/Year | Kelvin et al, 2019 (a) |
| --- | --- |
| Random sequence generation (selection bias) | **Unclear risk**  Randomized but method not specified. Distribution of baseline patient characteristics suggest randomization was achieved. |
| Allocation concealment  (selection bias) | **Low risk**  Participants were not informed they would be randomized to different arms so to avoid selection bias. |
| Blinding of participants and personnel (performance bias) | **High risk**  Participants nor personnel were blinded |
| Blinding of outcome assessment (detection bias) | **Unclear risk**  Unclear if outcome assessment came from self-report or facility records and if this was done equally across arms. |
| Incomplete outcome data  (attrition bias) | **Low risk**  No evidence of loss-to-follow-up |
| Selective reporting  (reporting bias) | **Low risk**  No evidence of selective reporting |
| Other bias | **Low risk** |

| Author/Year | Kelvin et al, 2019 (b) |
| --- | --- |
| Random sequence generation (selection bias) | **Unclear risk**  Randomized but method not specified. Distribution of baseline patient characteristics suggest randomization was achieved. |
| Allocation concealment  (selection bias) | **Low risk**  Participants were not informed they would be randomized to different arms so to avoid selection bias. |
| Blinding of participants and personnel (performance bias) | **High risk**  Participants nor personnel were blinded |
| Blinding of outcome assessment (detection bias) | **Unclear risk**  Unclear if outcome assessment came from self-report or facility records and if this was done equally across arms. |
| Incomplete outcome data  (attrition bias) | **Low risk**  No evidence of loss-to-follow-up |
| Selective reporting  (reporting bias) | **Low risk**  No evidence of selective reporting |
| Other bias | **Low risk** |

#### *4.2 QUADAS-2 Assessment for Diagnostic Accuracy*

Figure 4.2.1. Risk of bias graph: review of authors' judgements about each domain presented as percentages across included studies

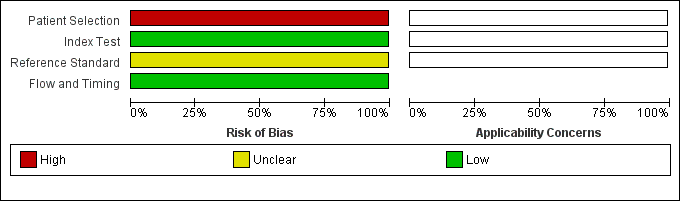

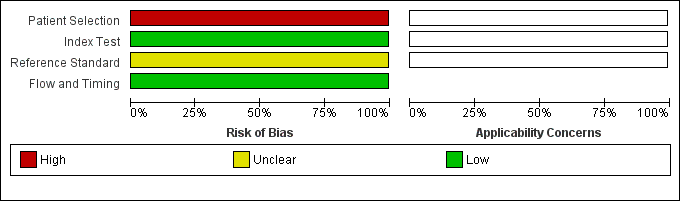

Figure 4.2.2. Risk of bias summary: review authors' judgements about each domain for each included study

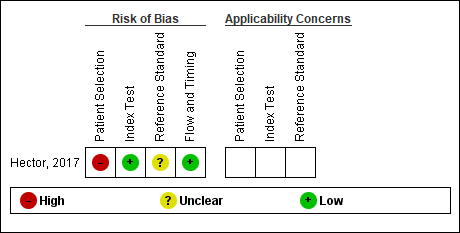

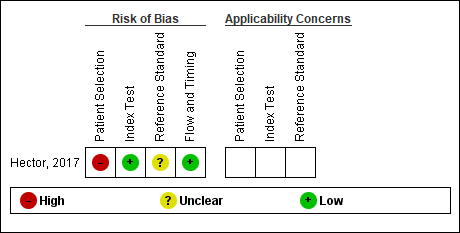

Table 4.2.1. QUADAS-2 Risk of bias table by study

| Author/Year | **Hector et al, 2017** |
| --- | --- |
| Patient selection | **High risk**  Participants self-selected, not randomized. |
| Index test | **Low risk**  Index test results were interpreted without knowledge of reference standard results. |
| Reference standard | **Unclear risk**  Reference standard results interpreted by a health-care worker with knowledge of results of index test. Reference standard included two tests and likely correctly classified results. |
| Flow and timing | **Low risk**  All patients received the same reference standard after index testing was conducted. |

#### *4.3 ROBINS-1 Assessment for observational studies*

Figure 4.3.1. Risk of bias graph: review of authors' judgements about each domain presented as percentages across included studies

|  | % |
| --- | --- |
| Confounding | 100 |
| Selection of participants | 100 |
| Classification | 100 |
| Deviations | 100 |
| Missing data | 100 |
| Outcomes measurement | 100 |
| Selective reporting | 100 |

Figure 4.3.2. Risk of bias summary: review authors' judgements about each domain for each included study

|  | Risk of Bias | | | | | | |
| --- | --- | --- | --- | --- | --- | --- | --- |
|  | Confounding | Selection of participants | Classification | Deviations | Missing data | Outcomes measurement | Selective reporting |
| Hector et al, 2017 | 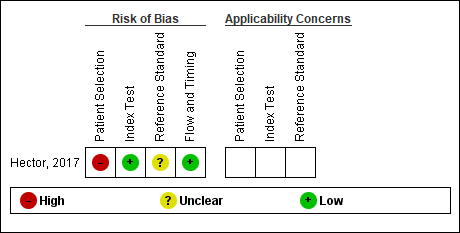 | 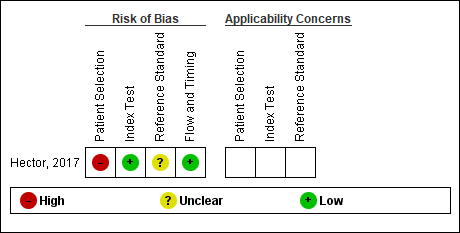 | 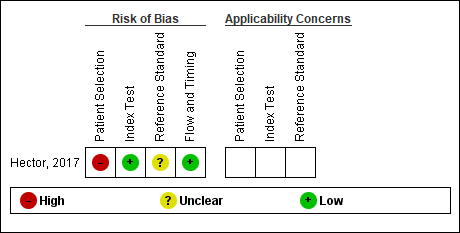 | 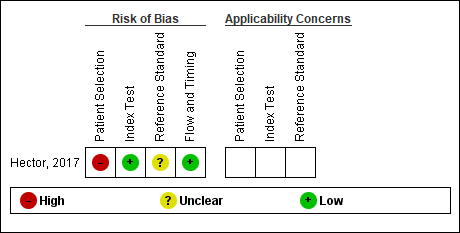 | 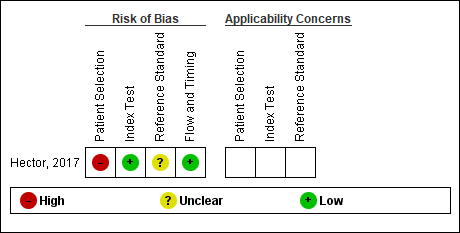 | 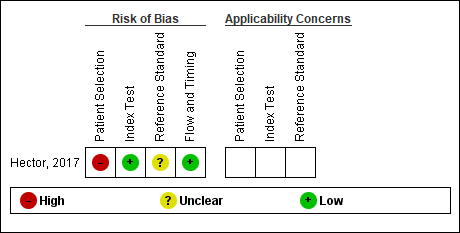 | 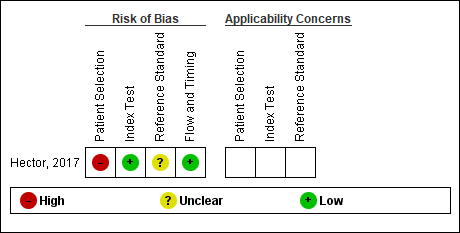 |
| Gaydos et al, 2013 | 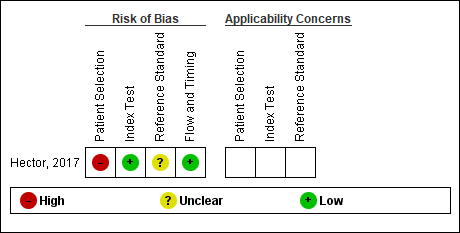 | 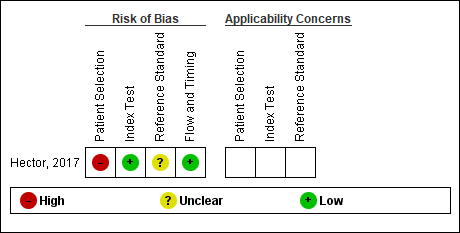 | 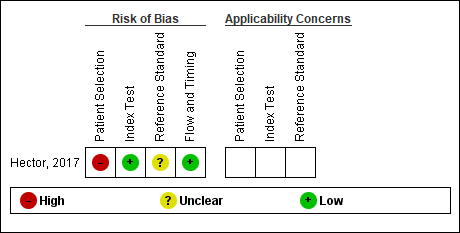 | 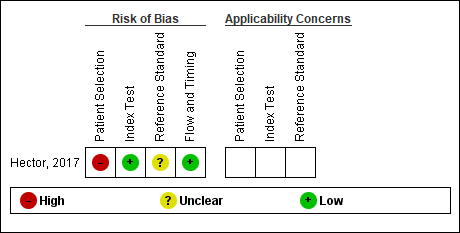 | 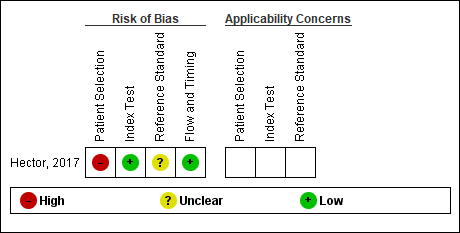 | 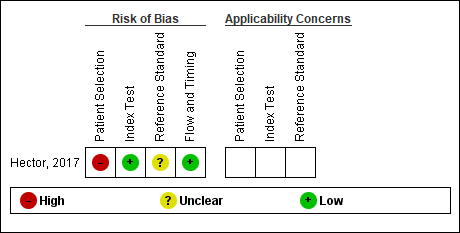 | 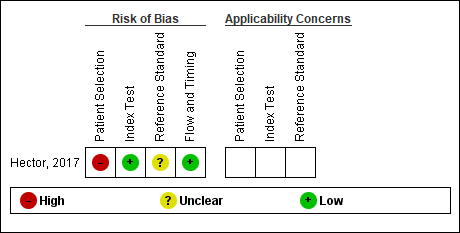 |

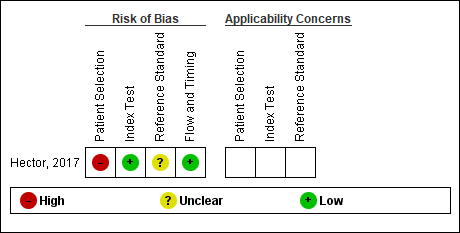

Table 4.3.1. ROBINS-1 Risk of bias table by study

| Author/Year | **Hector et al, 2017** |
| --- | --- |
| Bias due to confounding | **Critical**  Gender and race imbalance in participants demographic profile. |
| Bias in selection of participants | **Critical**  Participants self-selected into study after receiving sensitization at schools. Unknown how many participants consented to study after sensitization. Likely to have introduced a bias overestimating acceptability.  Furthermore, students have better access to sexual reproductive health education in comparison to out of school youth. Knowledge about HIV and testing uptake may be lower among out-of-school youth. |
| Bias in classification | **Low**  Single arm study trial |
| Bias due to deviations from intended interventions | **Low**  No evidence |
| Bias due to missing data | **Low**  No evidence |
| Bias in measurement of outcomes | **Critical**  Feasibility/ease of use is a self-reported outcome and is at high risk of social desirability bias |
| Bias in selection of the reported result | **Low**  No evidence |

| Author/Year | **Gaydos et al, 2013** |
| --- | --- |
| Bias due to confounding | **Critical**  Gender and race imbalance in participants demographic profile. |
| Bias in selection of participants | **Critical**  Participants self-selected into study. Among 955 patients approached, 49.5% consented to study. |
| Bias in classification | **Low**  Kiosk-facilitated and computer registered information |
| Bias due to deviations from intended interventions | **Low**  No evidence |
| Bias due to missing data | **Low**  No evidence |
| Bias in measurement of outcomes | **Critical**  Feasibility/ease of use is a self-reported outcome and is at high risk of social desirability bias |
| Bias in selection of the reported result | **Low**  No evidence |

### **Appendix C. GRADE table: Should HIV self-testing be offered as an additional test in health facilities?**

The following annex summarizes the certainty of evidence according to the GRADE approach. All outcomes in the GRADE table are presented in the order of criticalness determined by the guideline development group (GDG). Table 1 illustrates the full rankings of each outcome.

| **Certainty assessment** | | | | | | | **№ of patients** | | **Effect** | | **Certainty** | **Importance** |
| --- | --- | --- | --- | --- | --- | --- | --- | --- | --- | --- | --- | --- |
| **№ of studies** | **Study design** | **Risk of bias** | **Inconsistency** | **Indirectness** | **Imprecision** | **Other considerations** | **HIVST** | **SOC** | **Relative (95% CI)** | **Absolute (95% CI)** |  |  |
| **Uptake of HIV testing (all)** | | | | | | | | | | | | |
| 4^1,2,3,4^ | randomised trials | serious^a^ | not serious^b^ | not serious | serious^c^ | none | 687/2452 (28.0%) ^d^ | 261/2359 (11.1%) ^d^ | **RR 2.47** (0.96 to 6.33) | **163 more per 1,000** (from 4 fewer to 590 more) | ⨁⨁◯◯ Low | CRITICAL |
| **Uptake of HIV testing (compared to SOC)** | | | | | | | | | | | | |
| 4^1,2,3,4^ | randomised trials | serious^a^ | not serious^b^ | not serious | serious^c^ | none | 687/2452 (28.0%) ^d^ | 261/2359 (11.1%) ^d^ | **RR 2.47** (0.96 to 6.33) | **163 more per 1,000** (from 4 fewer to 590 more) | ⨁⨁◯◯ Low | CRITICAL |
| **Uptake of HIV testing (compared to enhanced SOC)** | | | | | | | | | | | | |
| 3^1,2,4^ | randomised trials | serious^e^ | not serious^f^ | not serious | not serious | none | 556/2302 (24.2%) | 156/2200 (7.1%) | **RR 3.17** (2.55 to 3.94) | **154 more per 1,000** (from 110 more to 208 more) | ⨁⨁⨁◯ Moderate | CRITICAL |
| **Uptake of HIV testing (general population)** | | | | | | | | | | | | |
| 1^4^ | randomised trials | very serious^g,h^ | not serious | not serious | not serious | none | 406/802 (50.6%) | 95/746 (12.7%) | **RR 3.98** (3.26 to 4.85) | **379 more per 1,000** (from 288 more to 490 more) | ⨁⨁◯◯ Low | CRITICAL |
| **Uptake of HIV testing (key and priority populations)** | | | | | | | | | | | | |
| 3^1,2,3^ | randomised trials | very serious^h,i^ | not serious^j^ | not serious | serious^k^ | none | 281/1650 (17.0%) | 166/1613 (10.3%) | **RR 2.07** (0.86 to 4.97) | **110 more per 1,000** (from 14 fewer to 409 more) | ⨁◯◯◯ Very low^l^ | CRITICAL |
| **Uptake of HIV testing (received sensitization)** | | | | | | | | | | | | |
| 3^1,2,4^ | randomised trials | very serious^h,m^ | not serious^n^ | not serious | not serious | none | 556/2302 (24.2%) | 148/2204 (6.7%) | **RR 3.27** (2.36 to 4.53) | **152 more per 1,000** (from 91 more to 237 more) | ⨁⨁◯◯ Low | CRITICAL |
| **Uptake of HIV testing (received no sensitization)** | | | | | | | | | | | | |
| 1^3^ | randomised trials | very serious^h,o^ | not serious | not serious | not serious | none | 131/150 (87.3%) | 113/155 (72.9%) | **RR 1.20** (1.07 to 1.34) | **146 more per 1,000** (from 51 more to 248 more) | ⨁⨁◯◯ Low | CRITICAL |
| **Uptake of HIV testing (in private booths)** | | | | | | | | | | | | |
| 1^4^ | randomised trials | very serious^g,h^ | not serious | not serious | not serious | none | 406/802 (50.6%) ^d^ | 95/746 (12.7%) ^d^ | **RR 3.98** (3.26 to 4.85) | **379 more per 1,000** (from 288 more to 490 more) | ⨁⨁◯◯ Low | CRITICAL |
| **Uptake of HIV testing (no private booths)** | | | | | | | | | | | | |
| 3^1,2,3^ | randomised trials | very serious^h,i^ | not serious^n^ | not serious | serious^p^ | none | 281/1650 (17.0%) | 166/1613 (10.3%) | **RR 2.07** (0.86 to 4.97) | **110 more per 1,000** (from 14 fewer to 409 more) | ⨁◯◯◯ Very low^l^ | CRITICAL |
| **Acceptability (would test again)** | | | | | | | | | | | | |
| 1^4^ | randomised trials | very serious^q^ | not serious | not serious | not serious | none | 399/402 (99.3%) ^r^ | 78/95 (82.1%) ^s^ | **RR 1.21** (1.10 to 1.33) | **172 more per 1,000** (from 82 more to 271 more) | ⨁⨁◯◯ Low | CRITICAL |
| **Acceptability (would recommend)** | | | | | | | | | | | | |
| 1^4^ | randomised trials | very serious^t^ | not serious | not serious | not serious | none | 403/406 (99.3%) ^u^ | 84/95 (88.4%) ^v^ | **RR 1.12** (1.04 to 1.21) | **106 more per 1,000** (from 35 more to 186 more) | ⨁⨁◯◯ Low | CRITICAL |
| **Acceptability (choice)** | | | | | | | | | | | | |
| 3^1,2,3^ | randomised trials | serious^i^ | not serious | not serious | not serious | none | Three randomized trials offered participants an option between 3 testing options. Among the participants enrolled in this arm, 16.78% (n=151/900) chose facility-based HIVST, 4% chose home-HIVST (n=36/900), and 10.33% (n=93/900) chose the standard of care.  ^w^ | | | | ⨁⨁⨁◯ Moderate | CRITICAL |
| **Accuracy (Ease of use)** | | | | | | | | | | | | |
| 2^5,6^ | observational studies | very serious^x^ | not serious | serious^y^ | not serious | none | Two cohort studies reported on ease of use. Pooled result found that among all interviewed participants, 75.33% (n=577/766) reported HIVST was easy to use compared to 2.87% (n=22/766) who reported HIVST was not easy to use. | | | | ⨁◯◯◯ Very low | CRITICAL |
| **Accuracy (spe, sen, ppv, npv)** | | | | | | | | | | | | |
| 1^5^ | observational studies | serious^z^ | not serious | not serious | serious^aa^ | none | One cohort study reported accuracy results. Out of 299 tests, there were 5 (1.67%) True Positives and 0 False Positives, 288 (96.32%) True Negatives and 0 False Negatives. There were 6 discrepancies, including 2 FB-HIVST interpreted as negative resulting in inconclusive confirmatory tests, and 1 FB-HIVST interpreted as invalid also resulted in an inconclusive confirmatory HIV test. Out of the 299 tests, 4 (1.3%) were therefore invalid. Excluding invalid/indeterminate results, specificity was measured at 1.00 [95% CI: 0.48, 1.00] and specificity at 1.00 [ 95% CI: 0.99, 1.00]. | | | | ⨁⨁◯◯ Low | CRITICAL |
| **Linkage to Care (among enrolled)** | | | | | | | | | | | | |
| 1^4^ | randomised trials | serious^ab^ | not serious | not serious | serious^ac^ | none | 7/802 (0.9%) ^d^ | 2/746 (0.3%) ^d^ | **RR 3.26** (0.68 to 15.62) | **6 more per 1,000** (from 1 fewer to 39 more) | ⨁⨁◯◯ Low | CRITICAL |
| **Linkage to Care (among tested positive)** | | | | | | | | | | | | |
| 1^4^ | randomised trials | very serious^ad^ | not serious | not serious | not serious | none | 19/28 (67.9%) ^ae^ | 5/6 (83.3%) ^ae^ | **RR 0.81** (0.52 to 1.26) | **158 fewer per 1,000** (from 400 fewer to 217 more) | ⨁⨁◯◯ Low | CRITICAL |
| **HIV Positivity (all, among enrolled)** | | | | | | | | | | | | |
| 4^1,2,3,4^ | randomised trials | serious^a^ | not serious^af^ | not serious | serious^ag^ | none | 21/2452 (0.9%) ^d^ | 4/2359 (0.2%) ^d^ | **RR 3.77** (0.81 to 17.44) | **5 more per 1,000** (from 0 fewer to 28 more) | ⨁⨁◯◯ Low | IMPORTANT |
| **HIV Positivity (compared to standard of care, among enrolled)** | | | | | | | | | | | | |
| 4^1,2,3,4^ | randomised trials | serious^a^ | not serious^af^ | not serious | serious^ag^ | none | 21/2452 (0.9%) | 4/2359 (0.2%) | **RR 3.77** (0.81 to 17.44) | **5 more per 1,000** (from 0 fewer to 28 more) | ⨁⨁◯◯ Low | IMPORTANT |
| **HIV Positivity (compared to enhanced standard of care, among enrolled)** | | | | | | | | | | | | |
| 3^1,2,4^ | randomised trials | serious^a^ | not serious | not serious | serious^ah^ | none | 21/2316 (0.9%) | 3/2213 (0.1%) | **RR 4.53** (1.54 to 13.34) | **5 more per 1,000** (from 1 more to 17 more) | ⨁⨁◯◯ Low | IMPORTANT |
| **HIV Positivity (general population, among enrolled)** | | | | | | | | | | | | |
| 1^4^ | randomised trials | very serious^a,h^ | not serious | not serious | serious^ai^ | none | 11/802 (1.4%) ^d^ | 2/746 (0.3%) ^d^ | **RR 5.12** (1.14 to 23.00) | **11 more per 1,000** (from 0 fewer to 59 more) | ⨁◯◯◯ Very low | IMPORTANT |
| **HIV Positivity (key & priority populations, among enrolled)** | | | | | | | | | | | | |
| 3^1,2,3^ | randomised trials | very serious^a,h^ | not serious^aj^ | not serious | serious^ah^ | none | 10/1650 (0.6%) | 2/1613 (0.1%) | **RR 2.97** (0.23 to 37.65) | **2 more per 1,000** (from 1 fewer to 45 more) | ⨁◯◯◯ Very low | IMPORTANT |
| **HIV positivity (received sensitization, among enrolled)** | | | | | | | | | | | | |
| 3^1,2,4^ | randomised trials | very serious^a,h^ | not serious | not serious | serious^ak^ | none | 21/2302 (0.9%) | 2/2204 (0.1%) | **RR 6.62** (1.97 to 22.24) | **5 more per 1,000** (from 1 more to 19 more) | ⨁◯◯◯ Very low | IMPORTANT |
| **HIV positivity (received no sensitization, among enrolled)** | | | | | | | | | | | | |
| 1^3^ | randomised trials | very serious^h,o^ | not serious | not serious | serious^al^ | none | 0/150 (0.0%) | 2/155 (1.3%) | **RR 0.21** (0.01 to 4.27) | **10 fewer per 1,000** (from 13 fewer to 42 more) | ⨁◯◯◯ Very low | IMPORTANT |
| **HIV Positivity (all, among tested)** | | | | | | | | | | | | |
| 4^1,2,3,4^ | randomised trials | very serious^am^ | not serious | not serious | serious^an^ | none | 21/687 (3.1%) ^d^ | 4/261 (1.5%) ^d^ | **RR 1.38** (0.45 to 4.19) | **6 more per 1,000** (from 8 fewer to 49 more) | ⨁◯◯◯ Very low | IMPORTANT |
| **Social harm (coerced to test)** | | | | | | | | | | | | |
| 1^4^ | randomised trials | very serious^ao^ | not serious | not serious | not serious | none | No participants who tested in the facility-based HIV self-testing arm reported coersion to test (n=0/1052). 4.03% (n=10/248) in the standard of care arm, and 3.82% (n=10/261) in the optimized standard of care arm reported coersion to test.  ^ap^ | | | | ⨁⨁◯◯ Low | IMPORTANT |
| **Social harm (coerced to disclose test results)** | | | | | | | | | | | | |
| 1^4^ | randomised trials | very serious^ao^ | not serious | not serious | not serious | none | No participants who tested in the facility-based HIV self-testing arm reported coersion to disclose test results (n=0/1052). One person (n=1/248) in the standard of care arm, and three persons (n=3/261) in the optimized standard of care arm reported coersion to disclose test results.  ^ap^ | | | | ⨁⨁◯◯ Low | IMPORTANT |

**CI:** confidence interval; **RR:** risk ratio

***Explanations***

a. Downgraded once. This was due to potential for performance bias (lack of blinding) in all trials, and high risk of detection bias (self-reported outcomes in Dovel, 2020) in 1 trial. Unclear risk of detection bias in 3 trials (Kelvin, 2018, 2019a, 2019b). Risk of selection bias was unclear in 3 trials (Kelvin, 2018, 2019a, 2019b). Unclear risk of reporting bias Dovel 2020 as HTS outcomes in partners specified in the trial registry was not reported in the manuscript.

b. Not downgraded. While statistical heterogeneity may be very high (Heterogeneity: Tau² = 0.88; Chi² = 187.21, df = 3, p < 0.00001 ; I² = 98%), subgroup analysis suggests heterogeneity may be explained by differences in populations (general vs. key populations) and whether participants received sensitization prior to intervention. Effects from individual and cluster RCTs consistently favoured FB-HIVST.

c. Downgraded once for wide 95% CI: 0.96-6.33

d. Results from one study were adjusted for cluster effect using study's reported intraclass correlation coefficient.

e. Downgraded once. This was due to potential for performance bias (lack of blinding) in all trials, and high risk of detection bias (self-reported outcomes in Dovel, 2020) in 1 trial. Unclear risk of detection bias in 3 trials (Kelvin 2019a, 2019b). Risk of selection bias was unclear in 3 trials (Kelvin 2019a, 2019b). Unclear risk of reporting bias in Dovel 2020 as HTS outcomes in partners specified in the trial registry was not reported in the manuscript.

f. Not downgraded. Statistical heterogeneity may be considered low (Heterogeneity: Tau² = 0.01; Chi² = 2.73, df = 2, p < 0.26 ; I² = 27%).

g. Downgraded once due to high risk of performance bias (lack of blinding), and high risk of detection bias (self-reported outcomes). Unclear risk of reporting bias Dovel 2020 as HTS outcomes in partners as specified in trial registry was not reported in the manuscript.

h. RoB is further downgraded as this is a sub-group analysis and the effect is no longer randomised.

i. Downgraded once. This was due to potential for performance bias (lack of blinding) in all 3 trials. There is also unclear risk of detection bias, and risk of selection bias.

j. While statistical heterogeneity may be very high (Heterogeneity: Tau² = 0.55; Chi² = 43.63, df = 2, p < 0.00001 ; I² = 95%), subgroup analysis suggests heterogeneity may be further explained by whether participants received sensitization prior to intervention or not. Effects from individual and cluster RCTs consistently favoured FB-HIVST.

k. Downgraded once for wide 95% CI: 0.86-4.97

l. Note certainty for this subgroup is very low because imprecision was downgraded one level

m. Downgraded once. This was due to potential for performance bias (lack of blinding) in all trials, and high risk of detection bias (self-reported outcomes in Dovel, 2020) in 1 trial. Unclear risk of detection bias in 2 trials, and unclear risk of selection bias in 2 trials. Unclear risk of reporting bias in Dovel 2020 as HTS outcomes in partners specified in the trial registry was not reported in the manuscript.

n. Not downgraded. Moderate statistical heterogeneity (Heterogeneity: Tau² = 0.05; Chi² = 4.96, df = 2, p < 0.08 ; I² = 60%), may further be explained by population types (general and key populations). Effects from individual and cluster RCTs consistently favoured FB-HIVST.

o. Downgraded once. This was due to potential for performance bias (lack of blinding). Unclear risk of detection bias, unclear risk of selection bias.

p. Downgraded once due to wide 95% CI: 0.86-4.97

q. Downgraded once due to high risk of performance bias (lack of blinding), and high risk of detection bias (self-reported outcomes) in 1 trial. Unclear risk of reporting bias Dovel 2020 as HTS outcomes in partners specified in the trial registry was not reported in the manuscript. Further downgraded because this reported outcome does not follow ITT analysis: denominator is not number enrolled into arm but number of participants tested, excluding those with previously known HIV-positive status.

r. Adjusted for cluster effect. Crude number of participants who reported they would test again: 1043/1052

s. Adjusted for cluster effect. Crude number of participants who reported they would test again: 204/248

t. Downgraded once due to high risk of performance bias (lack of blinding), and high risk of detection bias (self-reported outcomes) in 1 trial. Unclear risk of reporting bias Dovel 2020 as HTS outcomes in partners specified in the trial registry was not reported in the manuscript. Further downgraded because this reported outcome does not follow ITT analysis: denominator is not number enrolled into arm but number of participants tested.

u. Adjusted for cluster effect. Crude number of participants who reported they would recommend FB-HIVST to friends: 1054/1063

v. Adjusted for cluster effect. Crude number of participants who reported they would recommend PITC to friends: 219/248

w. No meta-analysis could be performed. Choice of different testing options was made within a single population.

x. Downgraded twice, due to high risk of confounding bias in both studies (gender, and race imbalance), high risk of participant selection in both studies (self-selected to participate in both studies, and school youth more likely to find test easy compared to out-of-school youth), high risk of attrition bias in 1 study (in Gaydos, among 955 patients approached 49.5% consented to study), high risk of bias for outcome measure as ease of use is a self-reported outcome.

y. Downgraded once due to differences in population (applicability): one study, conducted in 2012 in an inner-city US emergency department serving a disadvantaged population; other study, conducted in 2016 in two youth-friendly hospitals in Mozambique.

z. RoB using QADAS-2. Downgraded once due to high risk of bias for participant selection (self-selected group, not random), and because reference standard not blinded to index test results.

aa. Downgraded once due to large confidence interval for sensitivity (95% CI)= 0.48-1.00. Author notes that the study was not powered to calculate sensitivity and specificity of the oral HIV self-test, due to low positivity.

ab. Downgraded once. This was due to potential for performance bias (lack of blinding). Note that Detection bias is rated as low as linkage to care was both measured from self-reported data as well as data clerk review of clinical records at study facilities and surrounding facilities.

ac. Downgraded due to 1) large confidence interval and 2) because the confidence interval crosses the line of no effect.

ad. Downgraded once. This was due to potential for performance bias (lack of blinding). Note that Detection bias is rated as low as linkage to care was both measured from self-reported data as well as data clerk review of clinical records at study facilities and surrounding facilities. Downgrade another level because denominator here not those enrolled (ITT) but those tested positive. Not adjusted for cluster effect, but RoB downgraded already at lowest level.

ae. Not adjusted for cluster effect

af. Not downgraded as statistical heterogeneity may be considered low (Heterogeneity: Tau² = 0.88; Chi² = 4.67, df = 3, p < 0.20 ; I² = 36%) Heterogeneity is driven by one study, with non-significant results. Furthermore, heterogeneity may be explained by population differences: one study was conducted among general population, while remaining studies targeted key populations including truckers and female sex workers.

ag. Downgraded due to 1) very large confidence intervals, and 2) because 3 out 4 confidence intervals cross the line of no effect.

ah. Downgraded due to 1) large confidence intervals, and 2) because 3 out 3 confidence intervals cross the line of no effect.

ai. Downgraded once due to large confidence interval

aj. Not downgraded. While there is moderate heterogeneity (Heterogeneity: Tau² = 2.79; Chi² = 4.48, df = 2 (P = 0.11); I² = 55%), this is driven by one study, with non-significant results. Furthermore, heterogeneity may be explained by population differences: one study was conducted among general population, while remaining studies targeted key populations including truckers and female sex workers.

ak. Downgraded due to 1) large confidence intervals and 2) because 2 out 3 confidence intervals cross the line of no effect.

al. Downgraded because of large confidence interval and because the confidence interval crosses the line of no effect

am. Downgraded once due to potential for performance bias (lack of blinding) in all trials, and high risk of detection bias (self-reported outcomes in Dovel, 2020) in 1 trial. Unclear risk of detection bias in 3 trials (Kelvin, 2018, 2019a, 2019b). Risk of selection bias was unclear in 3 trials (Kelvin, 2018, 2019a, 2019b). Unclear risk of reporting bias Dovel 2020 as HTS outcomes in partners specified in the trial registry was not reported in the manuscript. Further downgraded because denominator here not ITT (those enrolled per arm) but number who tested for HIV.

an. Downgraded due to 1) large confidence intervals, and 2) because confidence intervals cross the line of no effect.

ao. Downgraded once due to high risk of performance bias (lack of blinding), and high risk of detection bias (self-reported outcomes) in 1 trial. Unclear risk of reporting bias Dovel 2020 as HTS outcomes in partners specified in the trial registry was not reported in the manuscript. Further downgraded because do not follow ITT measurement: denominator is not number enrolled into arm but number of participants tested, excluding those with previously known HIV-positive status. Not adjusted for cluster effect but RoB already at lowest level.

ap. As reported. Results not adjusted for cluster effect
